# Evidence for a role of skull bone marrow in human chronic pain

**DOI:** 10.1101/2025.07.19.25331817

**Authors:** Mehrbod Mohammadian, Nikos Efthimiou, Ludovica Brusaferri, Joya Cooper-Hohn, Minhae Kim, Jennifer P. Murphy, Zeynab Alshelh, Grace Grmek, Jack H. Schnieders, Courtney A. Chane, Thomas G. Carmichael, Danika Yang, Julia J. Schubert, Federico E. Turkheimer, Rafael A. Cazuza, Peter M. Grace, Ciprian Catana, Steven M. Stufflebeam, Robert R. Edwards, Vitaly Napadow, Kimberly Sullivan, Matthias Nahrendorf, Jodi M. Gilman, Marco L. Loggia

## Abstract

The skull bone marrow is emerging as a critical hub for neuroimmune signaling, yet its role in chronic pain is unknown. Using integrated positron emission tomography / magnetic resonance imaging, here we show that the levels of 18 kDa translocator (TSPO), a marker of immune cell density, is elevated in the skull bone marrow of individuals with chronic pain (N=125; chronic low back pain [cLBP], N=88; knee osteoarthritis [KOA], N=37), compared to healthy controls (N=22). These elevations were widespread, generally more pronounced for KOA than cLBP, and associated with greater pain intensity, pain interference, depression, and anxiety (all p < 0.001). Because TSPO is highly expressed in myeloid cells, these results associate skull bone marrow immune dysregulation with chronic pain and its associated psychological and functional impairments. These findings provide a strong rationale for investigating this previously overlooked structure, which remains largely underexplored in the context of pain.

## INTRODUCTION

Located within the diploë, the layer of trabecular bone enclosed between inner and outer tables within the skull, the skull bone marrow, is increasingly recognized as a source of immune cells in health and disease(*1–3*). Until recently, the central nervous system (CNS) was thought to be immunologically isolated from the rest of the body, and communication between the CNS and the immune system was thought to occur only under certain pathological conditions, when peripheral/circulating immune cells infiltrate the brain parenchyma after crossing the blood-brain barrier(*4*). However, this view has been challenged by the recent discovery of ossified vascular microchannels that traverse the inner skull cortex(*2*). These channels may serve as conduits for CNS-derived antigens, carried by the cerebrospinal fluid (CSF) from the brain into the skull marrow hematopoietic niche, where they signal and initiate myeloid egress. Conversely, myeloid cells can traffic from the marrow to the meninges and brain parenchyma, mediating inflammatory responses(*5*), as has been documented, for example, in rodent models following stroke or brain injury(*3*). Crucially, studies have shown that these myeloid cells have distinct immune signatures compared to the myeloid cells supplied from the blood, especially in pathological conditions that involve neuroinflammation(*5, 6*).

In a recent, preliminary report, we used integrated positron emission tomography/magnetic resonance imaging (PET/MRI) and the radioligand [^11^C]PBR28, to show that the levels of the 18 kDa translocator protein (TSPO) might be elevated in the skull bone marrow in individuals with migraine with visual aura(*1*). While TSPO is most commonly used as an imaging marker of brain inflammation (e.g., in Alzheimer’s disease(*7*), Parkinson’s disease(*8*), stroke(*9*), traumatic brain injury(*10*), chronic pain conditions(*11–14*)), it is also highly expressed in peripheral immune cells, making it a marker of immune cell density beyond the central nervous system(*15*). As such, the elevated TSPO PET signal we observed in the skull bone marrow in individuals with migraine suggests an increased density of myeloid cells in this compartment, implicating it as a potential contributor to the pathophysiology of this disorder. Notably, subsequent reports of similar findings in neurodegenerative and psychiatric diseases such as Alzheimer’s (*3*), stroke(*3*), depression(*16*), and autism(*17*), suggest that skull marrow activation – as measured in terms of TSPO PET signal elevation – is not unique to migraine, but may instead represent a more fundamental neuroimmune response shared across diverse neuroinflammatory conditions.

While the skull bone marrow is emerging as a site for neuroimmune signaling in various disorders, its involvement in chronic pain remains largely unknown. In this study, we aim to fill this gap by using TSPO PET to evaluate the role of skull bone marrow in adults with two different chronic pain conditions (i.e., chronic low back pain -cLBP- and knee osteoarthritis - KOA) and evaluate its association with the severity of pain and pain-comorbid symptoms. Given the emerging recognition of the skull bone marrow as a potentially critical hub for neuroimmune signaling, unveiling its involvement in chronic pain could pave the way for the investigation of novel mechanisms underlying persistent pain, with implications for the development of novel diagnostic and therapeutic tools.

## RESULTS

### Demographics and clinical characteristics

Demographic and baseline clinical data are summarized in Table 1.

**Table 1.** Participant Demographics by Study Group.

| Variable | cLBP | KOA | cLBP + KOA<br>“Pain group” | HC |
| --- | --- | --- | --- | --- |
| N | 88 | 37 | 125 | 22 |
| Female, N(%) | 52 (59.09) | 16 (43.24) | 68 (54.4) | 11 (50) |
| Age, mean (SD) | 47.32 (15.45)<sup>*†</sup> | 66.59 (8.29)<sup>*</sup> | 53.1 (16.38) | 55.68 (16.89) |
| TSPO Genotype, N(%HAB) | 54 (61.36) | 21(56.76) | 75 (60) | 10 (45.45) |
| PROMIS-29 [0-20, except where specified], mean (SD) |  |  |  |  |
| Pain intensity [0-10] ↑ | 4.17 (1.79)<sup>**</sup> | 4.32 (2.54)<sup>**</sup> | 4.22 (2.03)<sup>**</sup> | 0.36 (0.66) |
| Pain interference ↑ | 8.85 (3.21)<sup>**†</sup> | 11.08 (4.15)<sup>**</sup> | 9.51 (3.64)<sup>**</sup> | 4.00 (0) |
| Physical function ↓ | 12.64 (5.88)<sup>**</sup> | 13.40 (3.65)<sup>**</sup> | 12.86 (5.32)<sup>**</sup> | 19.95 (0.21) |
| Anxiety ↑ | 6.79 (3.37)<sup>*</sup> | 6.27 (2.13) | 6.64 (3.05)<sup>*</sup> | 5.22 (1.90) |
| Depression ↑ | 5.65 (2.59)<sup>*</sup> | 4.97 (1.40) | 5.45 (2.32)<sup>*</sup> | 4.64 (1.29) |
| Fatigue ↑ | 9.06 (3.43)<sup>**</sup> | 8.19 (2.56)<sup>**</sup> | 8.80 (3.21)<sup>**</sup> | 6.04 (1.84) |
| Sleep disturbance ↑ | 10.34 (3.67)<sup>**</sup> | 10.08 (3.37)<sup>**</sup> | 10.26 (3.57)<sup>**</sup> | 6.86 (2.75) |
| Social roles ↓ | 12.11 (5.41)<sup>**</sup> | 10.16 (4.15)<sup>**</sup> | 11.54 (5.13)<sup>**</sup> | 18.14 (2.47) |
| KOA, knee osteoarthritis; cLBP, chronic low back pain; HC, healthy controls; TSPO, 18 kDa translocator protein; HAB, high-affinity binder; SD, standard deviation; |  |  |  |  |
| Vs HC: <sup>*</sup>p<0.05, <sup>**</sup>p<0.001; cLBP Vs. KOA: <sup>†</sup>p<0.05; ↑/↓, directionality of worse outcome |  |  |  |  |

The Pain group comprised individuals with knee osteoarthritis (KOA, N=37) or chronic low back pain (cLBP, N=88). Pain and HC groups did not significantly differ in sex, age, or Ala147Thr TSPO genotype distribution (which influences the binding affinity of the radioligand [^11^C]PBR28(*18, 19*)). Although not statistically significant, the Pain group tended to have a larger body weight than the HC group ([individuals with chronic pain: 80.49 ± 16.43]; [healthy controls: 73.75 ± 12.96], p = 0.07).

As expected, the Pain group reported on average higher levels of pain intensity (4.22 ± 2.03 on a 0 to 10 scale) and pain interference (9.51 ± 3.64 out of 20) compared to the HC group. Among participants within the Pain group who were able to quantify the duration of their pain (N=110), the average reported duration was 7.82 ± 8.40 years overall, with similar values observed in both the cLBP (7.67 ± 7.80) and KOA groups (8.22 ± 9.57). Additionally, individuals in the Pain group reported on average higher levels of anxiety (6.64 ± 3.05), depression (5.45 ± 2.32), fatigue (8.8 ± 3.21), and sleep disturbance (10.26 ± 3.57), but lower (i.e., worse) scores on physical function (12.86 ± 5.32) and social roles (11.54 ± 5.13), compared to the HC group (p’s < 0.05).

Converting the raw PROMIS-29 scores to T-scores revealed substantial interindividual variability in the severity levels, with most participants within the Pain group endorsing mild symptoms (55≤T-score<60 for most domains; 40≤T-score<45 for social roles and physical function), but several reporting moderate (60≤T-score<70 and 30≤T-score<40, respectively) or even severe (T-score≥70 and <30, respectively). As expected, the scores from the healthy controls were generally mild and limited in range (and thus associations between imaging and clinical variables were not explored in this group). See *Supplementary Results* and Fig. S1 for the distribution of the T-scores across all domains for all participants, including comparisons of demographic and clinical characteristics between patient subgroups.

### TSPO PET signal in relation to age and sex

In preparation to our main imaging analyses, we first examined the relationship between skull bone marrow TSPO signal and both sex and age to determine whether these variables should be included as covariates in our statistical models. In these analyses, the main effects of these demographic variables on the PET signal were computed by modeling the HC group and the two Pain subgroups (cLBP and KOA) as separate regressors, and then averaging the resulting contrasts to derive a single overall estimate.

In these analyses, the PET signal was negatively correlated with age, with significant, and very widespread, clusters spanning bilaterally across the parietal and frontal bones (Fig. S2A). When plotted separately for HC and Pain subgroups, the average TSPO PET signal from a 5mm-radius sphere centered on the voxel of peak significance (MNI coordinates: x=139, y=99, z=140) and constrained by the study-specific skull mask (See Methods) revealed comparable age-related PET signal decreases across all groups (Fig. S2B). Additionally, female participants exhibited some regions of the skull bone marrow with higher TSPO PET signal than males, although this sex effect was limited to a relatively small area within the medial occipital region of the skull (Fig. S2C). Similarly to the age results, plotting the average signal around the peak significance (MNI coordinates: x=91, y=24, z=88) revealed comparable sex effects in all groups (including both Pain subgroups when examined independently) (Fig. S2D). For illustrative purposes, average skull TSPO PET maps stratified by age tertiles and sex are shown in Fig. S3. Given the established associations between TSPO PET signal and both age and sex, these variables were included as covariates in all subsequent analyses.

### Group differences in TSPO PET signal

The overall spatial distribution of the TSPO PET signal was comparable across groups, with both Pain and HC groups showing similar areas of peak intensity in the lateral part of the frontal bone, the dorsomedial portion of the parietal and occipital bones, and the rostral and caudal portions of the parietal skull (Fig. 1A). While both groups showed a similar spatial pattern, the Pain group (derived by modeling KOA and cLBP as separate regressors, and then averaging the two contrasts to derive a single group estimate) exhibited markedly higher TSPO PET signal across large portions of the skull compared to HC, with particularly pronounced increases observed bilaterally in the frontal and parietal regions (Fig. 2B). A follow-up analysis modeled KOA and cLBP subgroups as separate groups, focusing on the area of peak significance in the Pain>HC contrast (MNI coordinates: x=117, y=196, z=98). This analysis revealed that 1) both cLBP and KOA subgroups demonstrated a statistically significant increase in skull TSPO PET signal compared to healthy controls and that 2) KOA demonstrated the highest signal (Fig. 1C, also see Supplementary Results and Fig. S4). Additionally, there were no significant effects of sex, or group-by-sex interactions (Fig. 1D), consistent with the minimal overlap between the cluster significant in the Pain>HC contrast and the cluster exhibiting a sex effect (Fig. S2C).

**Fig. 1.**
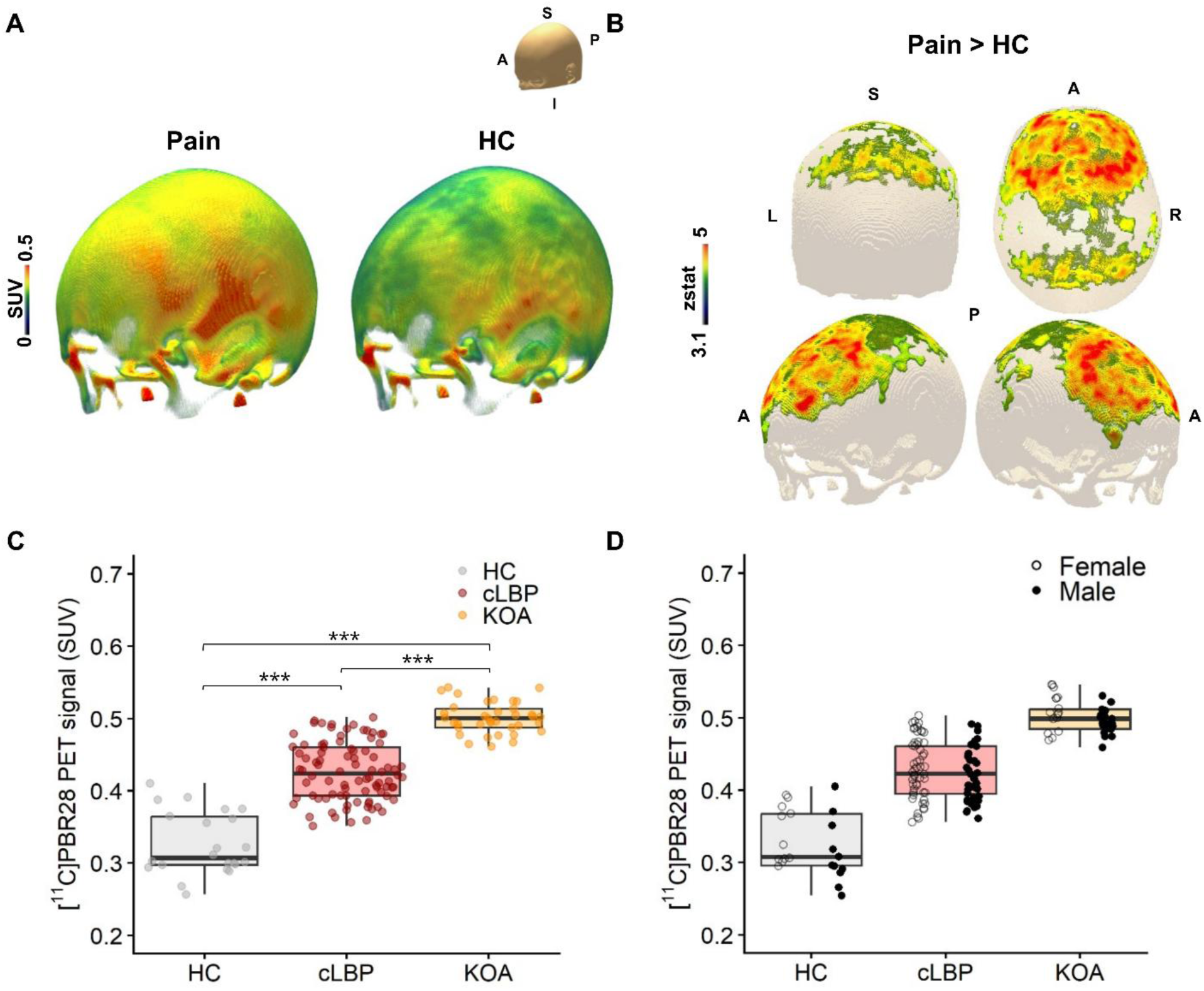
Skull TSPO PET differences between pain and control groups. Average [11C]PBR28 TSPO PET skull maps (SUV) for both Pain (averaging all individuals with KOA or cLBP) and HC groups (**A**); 3D rendering of the statistical contrast showing elevated TSPO PET SUV in the Pain group relative to HC, computed in the standard MNI152 space (**B**); Box and scatter plots depicting mean TSPO PET SUV from the skull region around the peak significance (See Methods) (**C**); Box and scatter plots showing mean signal from the skull region around the peak significance, shown separately for each group and stratified by sex (**D**). SUV: Standardized Uptake Value.

**Fig. 2.**
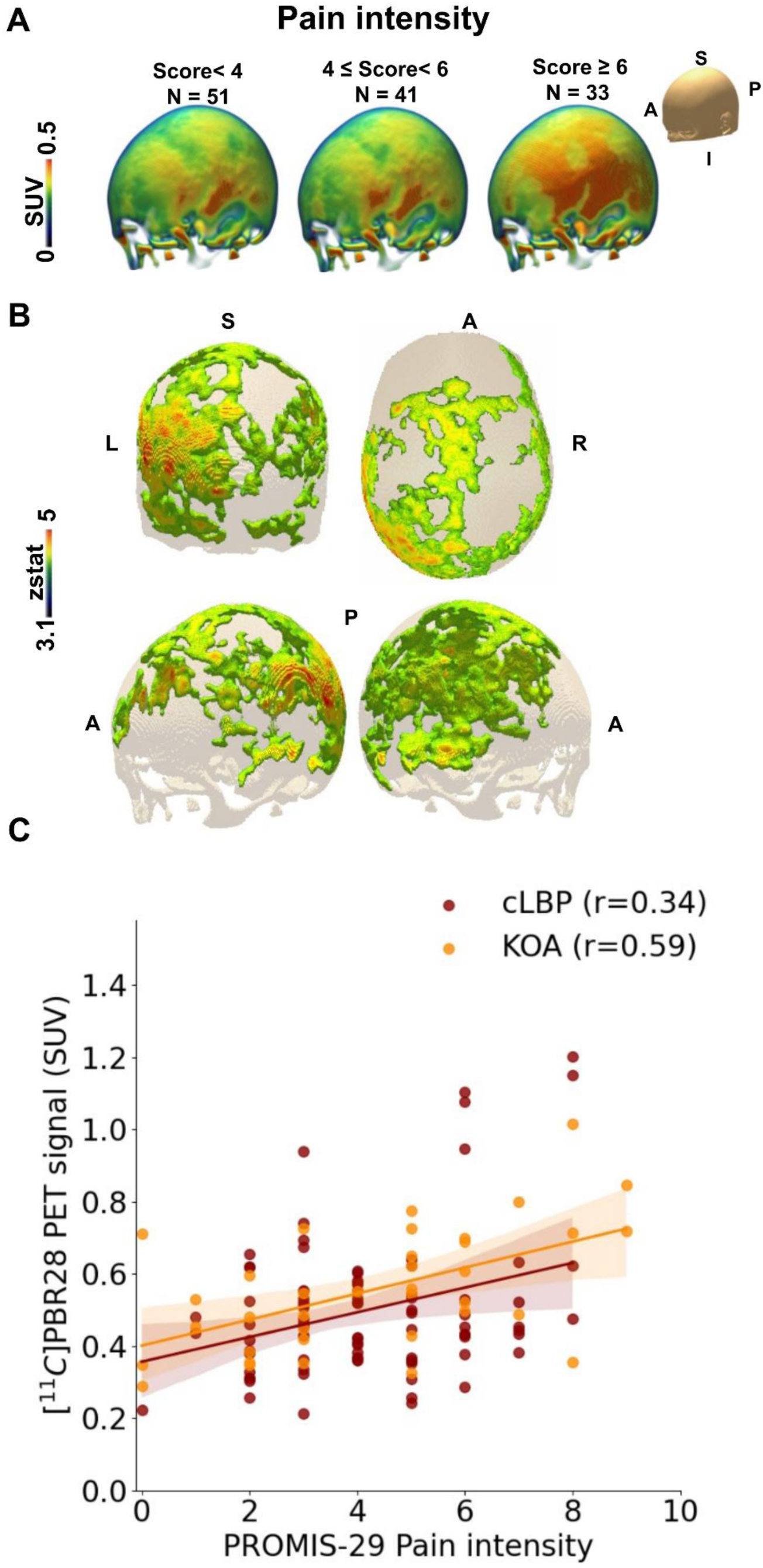
Skull TSPO PET association with pain intensity. Average [11C]PBR28 TSPO PET skull SUV maps for participants in Pain group, divided into low, mid, and high tertiles based on pain intensity scores (**A**); 3D rendering of the statistical contrast showing a positive correlation between the TSPO PET SUV and the PROMIS-29 pain intensity score (**B**); Scatter plots showing the mean signal averaged across significant voxels surrounding the peak, with fitted regression lines and 95% confidence intervals, presented separately for each Pain subgroup. (**C**).

### Association with pain-related scores

To understand the clinical significance of the group differences in skull PET signal, we evaluated the association between imaging and clinical variables in the Pain group. Pain intensity and pain interference were positively and significantly correlated with skull TSPO PET signal in the Pain group. For both variables, the effects were widespread, spanning temporal, parietal, and occipital portions of the skull. The association with pain intensity was most prominent and widespread, with larger involvement of the frontal portions of the skull, and showed some left-sided dominance, with frontal, parietal, temporal, and occipital peaks being more prominent on the left hemisphere (Fig. 2). The association between the TSPO PET signal and pain interference was less widespread but also showed some left-sided dominance, particularly in the caudal part of the parieto-occipital portion of the skull (Fig. 3). Once again, the average signal extracted from the region of peak significance in these analyses (MNI coordinates: pain intensity, x=159, y=64, z=83; pain interference, x=113, y=28, z=40) demonstrated similar patterns in both Pain subgroups independently (Figs. 2C, 3C).

**Fig. 3.**
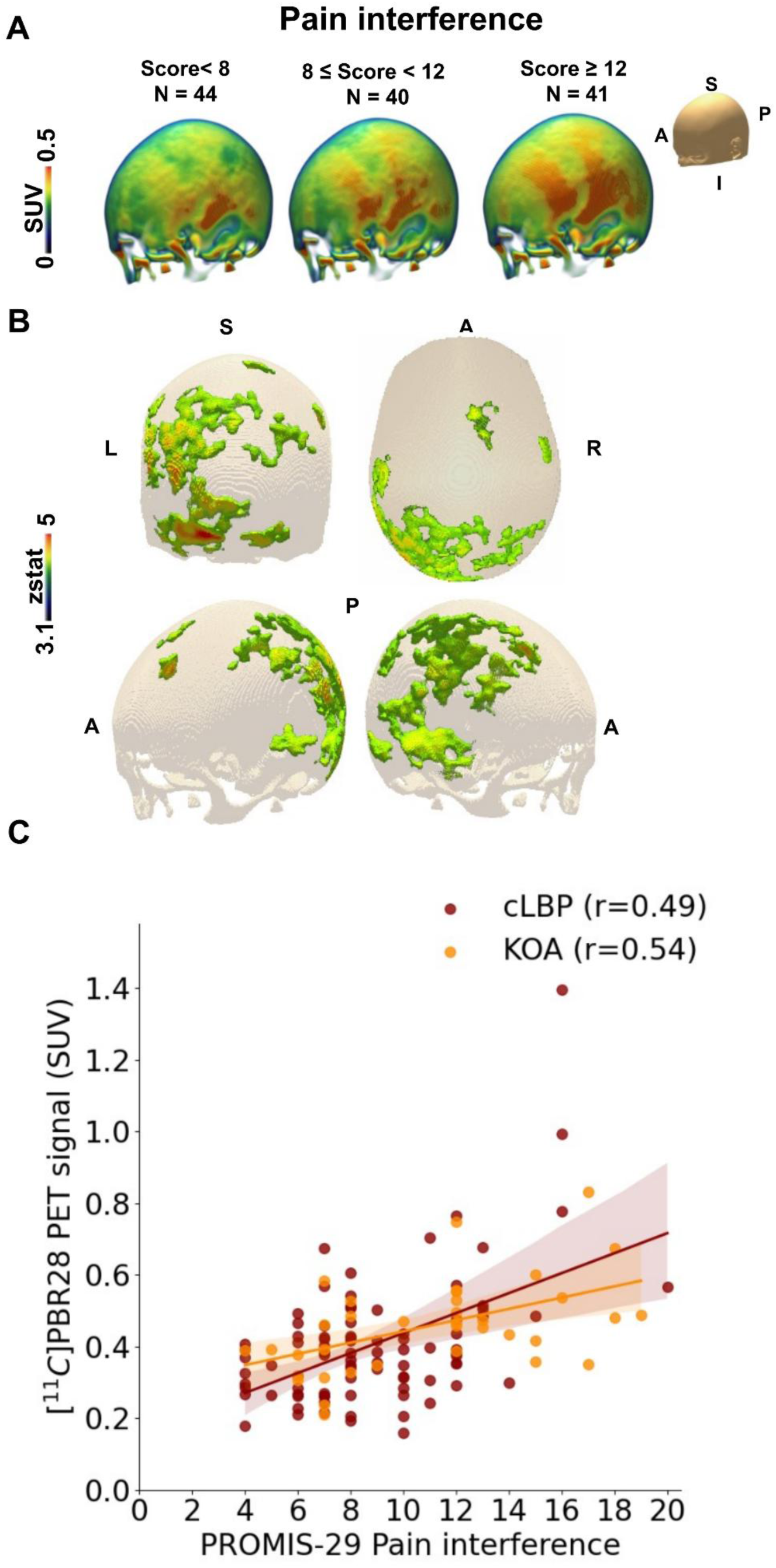
Skull TSPO PET association with pain interference. Average [11C]PBR28 TSPO PET skull SUV maps for participants in Pain group, divided into low, mid, and high tertiles based on pain interference scores (**A**); 3D rendering of the statistical contrast showing a positive correlation between the TSPO PET SUV and the PROMIS-29 pain interference score (**B**); Scatter plots showing the mean signal averaged across significant voxels surrounding the peak, with fitted regression lines and 95% confidence intervals, presented separately for each Pain subgroup. (**C**).

### Association with other patient-reported scores

To further understand the clinical significance of the skull TSPO PET signal, we evaluated its association with the other domains of the PROMIS-29 questionnaire. Across the entire Pain group, we observed a statistically significant positive correlation with the mental health domains of the PROMIS-29 questionnaire, namely anxiety (Fig. 4A-B) and depression (Fig. 4C-D). These associations were primarily localized over the dorsomedial portions of the frontal and parietal bones, with the anxiety analysis yielding a slightly more extended cluster (Fig. 4A) compared to the depression analysis (Fig. 4C). Once again, average signals extracted from the regions around the peak significance (MNI coordinates: anxiety, x=87, y=156, z=146; depression, x=86, y=121, z=160) revealed similar associations in both KOA and cLBP independently (Figs. 4B, 4D).

**Fig. 4.**
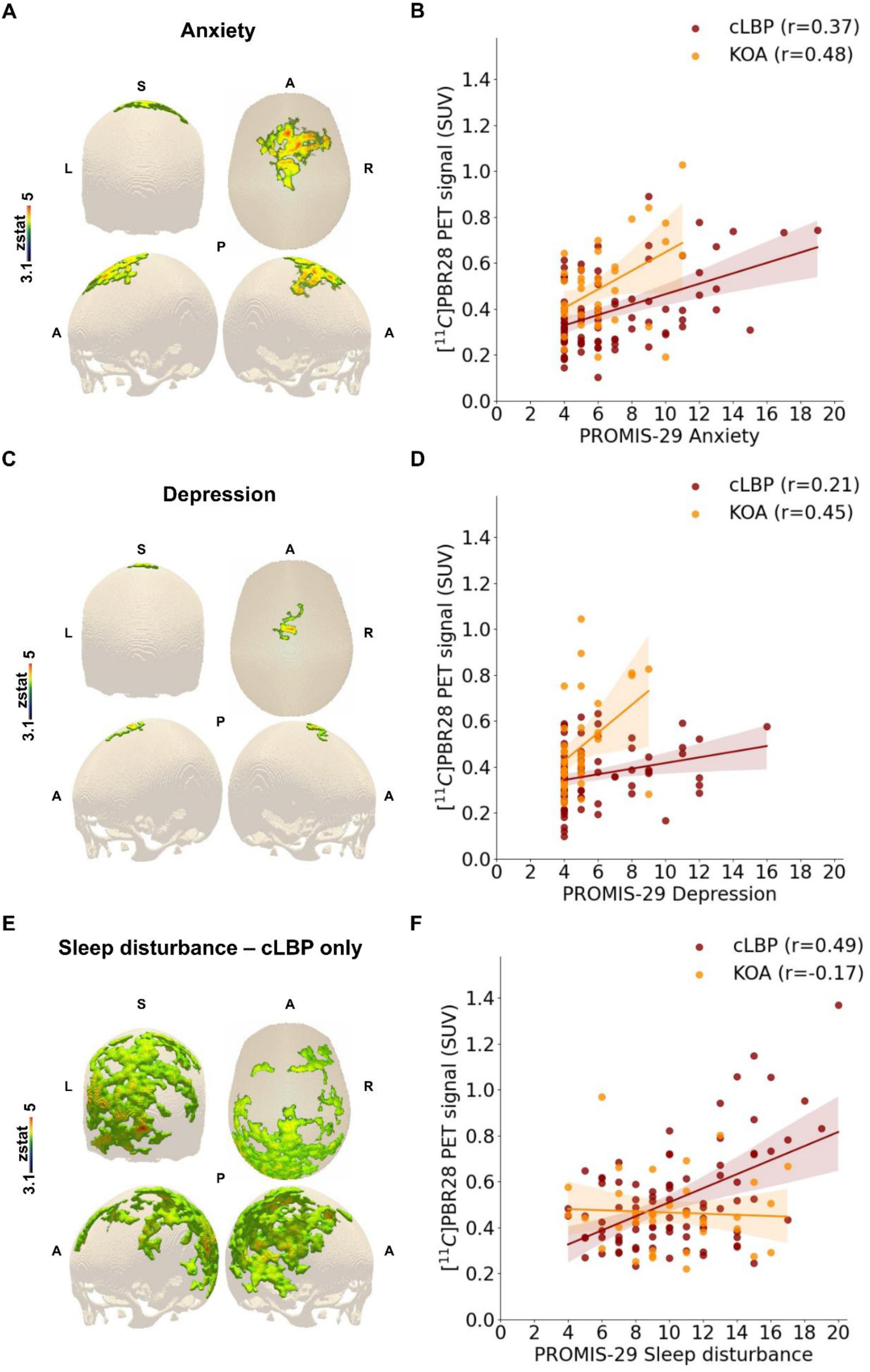
Skull TSPO PET associations with non-pain related PROMIS 29 scores. 3D rendering, and relative scatterplots, of the statistical contrasts showing positive correlations between the TSPO PET SUV and the PROMIS-29 anxiety (Pain group; **A-B**), depression (Pain group; **C-D**), and sleep disturbance (cLBP only; **E-F**) scores;

No significant correlations were observed between the TSPO PET signal and other PROMIS-29 domains across the Pain group as a whole. However, an association was observed between sleep disturbance and skull TSPO PET signal in cLBP individuals, spanning fronto-parietal and parieto-occipital regions with a left side dominance (Fig. 4E). Extraction of the average signal from the peak significant voxel (MNI coordinates: x=92, y=16, z=65) confirmed this association in the cLBP subgroup only (Fig. 4F).

## DISCUSSION

An emerging body of research(*1, 3, 16, 17*) identifies the skull bone marrow as a potential key structure in the pathophysiology of many conditions, with possible implications for the development of new biomarkers and therapeutic targets for a broad range of human chronic conditions. Building on these findings, here we present evidence supporting a role for skull TSPO PET signal in human chronic pain conditions, leveraging a large patient dataset (N=125).

While the specific cellular mechanisms underlying our imaging results remain to be elucidated, one potential explanation for the elevated TSPO signal in the skull is neuroimmune crosstalk between the brain and skull bone marrow, mediated by recently identified vascular channels that directly connect these regions(*2, 20*). These channels may serve as conduits for 1) CNS-derived antigens, carried by the cerebrospinal fluid from the brain into the skull marrow hematopoietic niche, where they signal and initiate myeloid egress, and 2) myeloid cells trafficking from the marrow to the meninges and brain, to mediate a neuroinflammatory response. This response has been documented, for instance, in rodents after a stroke or brain injury(*3*). Recent evidence suggests that neutrophils may play a key role underlying these mechanisms. Kolabas et al. reported significantly higher TSPO RNA expression in the skull bone marrow of injured versus naïve mice, particularly within neutrophil populations(*3*). This raises the possibility that neutrophils contribute to the skull TSPO PET signal observed in our studies, potentially reflecting an increased cellular density. Furthermore, our previous work has demonstrated elevated TSPO PET signal in the brains of individuals with various chronic pain conditions, including KOA and cLBP(*12, 13, 21*). We speculate that inflammatory signals originating in the brain may propagate through the skull–meninges channels, thereby activating resident skull marrow immune cells and promoting the mobilization of myeloid cells, including neutrophils. This brain inflammation may, in turn, be sustained by persistent nociceptive input from peripheral pathology (e.g., in the joint or spine), consistent with the concept of ‘neurogenic neuroinflammation’ (*22*).

Remarkably, we found a statistically significant and positive correlation between the TSPO PET signal in the skull and the severity of pain and pain-comorbid symptoms, highlighting the potential clinical relevance of our observations. When KOA and cLBP contrasts were averaged to derive a single Pain group estimate, elevated PET signal was associated with higher pain intensity and interference, as well as anxiety and depression. Importantly, follow-up desegregated analyses revealed that comparable effects could be replicated within each Pain subgroup despite the different pain etiologies (back or knee pain), indicating remarkable generalizability. Interestingly, the KOA group appeared to have the highest skull TSPO PET signal, followed by the cLBP and healthy control groups, for reasons that remain to be elucidated. Additionally, the distinct spatial distribution of the skull TSPO PET signal observed for different symptoms (i.e., anxiety, depression, and pain intensity/interference) suggests a possible disease- or state-specific topographical signature that warrants further examination. Future studies are needed to determine whether distinct portions of the skull bone marrow, perhaps owing to their proximity to different brain structures, might serve specialized functions.

Notably, a significant negative correlation was observed between skull TSPO signal and age in both healthy controls and individuals with chronic pain. Similar findings were recently reported in individuals with Alzheimer’s disease(*3*), further underscoring the generalizability of this observation. This result is somewhat counterintuitive, as aging is generally thought to be accompanied by an increase in inflammation within the brain. However, recent evidence suggests that skull bone marrow undergoes continuous expansion in both volume and vascularization throughout adulthood(*6, 23*). Intriguingly, the rate of such decline in skull TSPO signal appeared qualitatively steeper for the chronic pain groups. Nevertheless, future studies with larger samples of healthy controls might be needed to test this hypothesis formally.

In a dorsomedial portion of the skull, we observed elevated TSPO PET signal female subjects compared with males. This sex-related difference is broadly consistent with prior evidence indicating that neuroimmune activity differs inherently between sexes(*24*). Such sexual dimorphism in immune–pain interactions has been proposed to contribute to differential susceptibility to chronic pain conditions and to distinct neuroinflammatory signaling pathways across sexes. Notably, however, the dorsomedial skull region showing higher TSPO binding in females did not spatially overlap with regions that displayed increased TSPO signal in the Pain group relative to healthy controls, suggesting that these effects likely reflect independent biological mechanisms rather than a shared pain-related process.

The present study has several notable strengths, including a large sample size and the inclusion of two separate groups of individuals with chronic pain. However, there are several limitations. Perhaps most importantly, in this cross-sectional analysis, it is impossible to determine whether the TSPO PET signal is caused by the painful condition or precedes it. Future longitudinal work assessing skull TSPO PET signal before and after the establishment of a chronic pain condition (e.g., due to a surgery with a relatively high likelihood of chronic post-surgical pain, and/or within the context of a preclinical pain model) will be needed to answer the question of causality. Also crucially, the focus of the current manuscript was not an analysis of the relationship between skull and brain inflammation, as a single metric suitable for quantifying the PET signal for both regions remains to be identified. Indeed, SUV is generally accepted for peripheral structures, but less so for the quantification of the brain.

In conclusion, these findings indicate that increased immune activity in the skull bone marrow is linked to chronic pain and its associated symptoms. This highlights the emerging importance of skull bone marrow immunity in the development of chronic pain and possibly other conditions involving brain inflammation. Overall, these insights position the skull bone marrow as a crucial player in neuroimmune responses, with immune cell density in this area serving as a potential biomarker and therapeutic target for brain inflammation. These findings provide a strong rationale for investigating this previously overlooked structure, which remains largely underexplored in the context of pain.

## MATERIALS AND METHODS

### Participants

A total of 125 adults with chronic pain (age, mean ± standard deviation: 53.10 ± 16.38 years old) were included in this study. Of these, 88 were individuals with cLBP (47.32 ± 15.45 years old), recruited from two clinical trials (NCT03106740 and NCT03891264), and 37 were individuals with KOA (66.59 ± 8.29 years old) who were scheduled to undergo total knee replacement surgery. The present study included only data collected prior to the initiation of interventions (pharmacological or surgical). In addition, data from 22 healthy controls (55.68 ± 16.89 years old) were included in these analyses. All participants were adults, aged 18 years or older, and provided written informed consent for their inclusion in the respective research studies. Detailed inclusion and exclusion criteria are available in recent publications from our group regarding these patient cohorts(*21, 25*). All participants were genotyped for the Ala147Thr polymorphism in the *TSPO* gene, which predicts binding affinity of [^11^C]PBR28(*18, 19*) for its target, and only individuals with the Ala/Ala or Ala/Thr genotypes (corresponding to high-affinity and mixed-affinity binders, respectively) were included.

### TSPO PET-MR imaging and image analysis

All participants underwent integrated PET/MRI sessions of 90-minute brain [^11^C]PBR28 PET scan using a Siemens Biograph mMR scanner (Siemens Healthineers, Erlangen, Germany). A maximum of 15 mCi ([Pain group: 14.08 ± 1.32 mCi]; [HC group: 14.01 ± 1.73 mCi], p = 0.83) of [^11^C]PBR28 was injected intravenously with a slow bolus over a 30–60-second period by a licensed nuclear medicine technologist. Dynamic PET scans were acquired in list mode, and static reconstructions were generated from the PET data 60-90 minutes after radiotracer injection. Image reconstruction was performed with an in-house developed framework for Correction of Motion and Brain Alignment (COMBRA)(*26*) using ordered subset expectation maximization (OSEM)(*27*) with 21 subsets and 3 iterations, as implemented in Siemens e7 tools. Corrections for background, frame duration, and decay were applied in accordance with the manufacturer’s guidelines. Alongside PET data collection, MR images were acquired, including a multi-echo T1-weighted MPRAGE sequence (TR/TE1/TE2/TE3/TE4 = 2530/1.69/3.55/5.41/7.27 ms, flip angle = 7°, voxel size = 1 mm³). These images were used for anatomical reference and to create a m map (“pseudoCT”) used for attenuation correction(*28*), as well as to create skull masks (see below). While previous studies have already shown an overall good agreement between pseudoCT and CT(*29, 30*), we performed a further, skull-focused, validation in a separate group of participants who underwent both PET/MRI and PET/CT (see Supplementary Methods). In these participants, skull PET uptake obtained using an MR-based pseudoCT- or CT-based attenuation correction yielded very comparable results (see Supplementary Results), supporting the validity of our approach.

PET signal was quantified as the Standardized Uptake Value (SUV), calculated as the mean radioactivity concentration divided by the ratio of the administered dose to body weight.

Fig. 5 illustrates the general workflow of the analysis pipeline. PseudoCT images, were generated from the T1-weighted MPRAGE scans(*28*). As opposed to prior work that adopted on region-of-interest analysis focused on discrete skull regions(*1*), here we developed a pipeline to examine the entire skull using a voxel-wise approach, enabling a more comprehensive characterization of skull bone marrow and its relationships with symptom severity. A skull mask was created by applying a heuristic threshold [0.13] to the pseudoCT image to isolate bone values. Any gaps between inner and outer skull layers or other inaccuracies resulting from this thresholding process were corrected by manual editing. An affine transformation matrix was calculated to co-register the PET SUV images to the structural T1-weighted image. The TSPO PET SUV image for each subject was then masked with the skull mask to produce an estimated skull PET SUV image. Additionally, to reduce the possibility of contamination from brain signal in the skull PET data, all T1-weighted MR images were processed using the FreeSurfer(*31*) ‘recon-all’ pipeline, and the resulting brain mask was used to subtract the brain from the estimated skull PET SUV image.

**Fig. 5.**
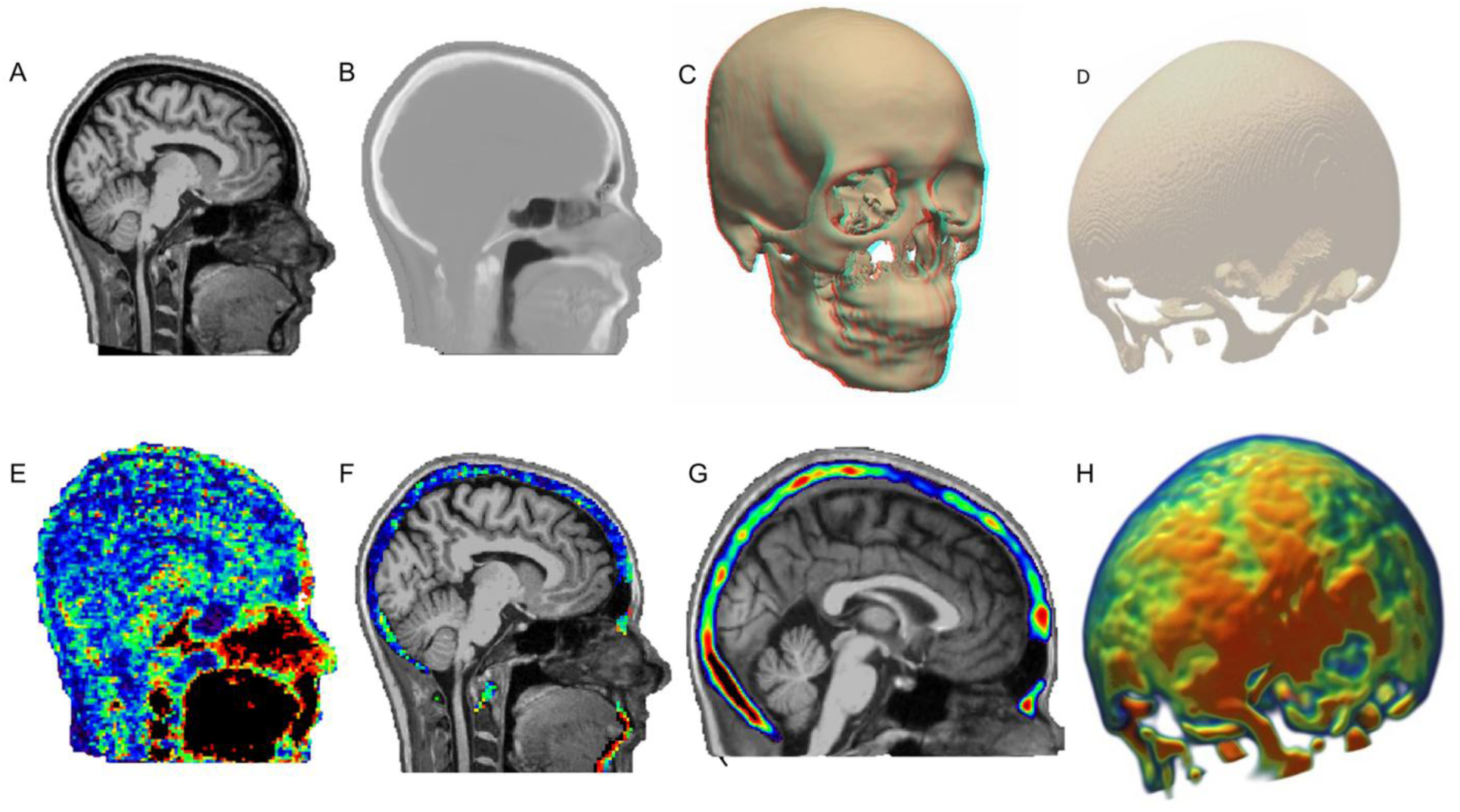
Processing pipeline (representative subject). T1-weighted structural MRI (**A**); Estimated pseudoCT image used for PET attenuation correction (**B**); Reconstructed 3D rendering of a subject’s skull in native space after applying the heuristic threshold to extract the bone values (**C**), highlighting the pseudoCT method’s ability to accurately recover the skull shape from the T1-weighted MRI, which is typically unable to visualize this structure; Reconstructed 3D rendering of the skull in standard MNI152 space (**D**); Reconstructed average 60-90 minute [11C]PBR28 TSPO PET SUV (**E**); Unsmoothed skull [11C]PBR28 TSPO PET SUV overlaid on structural T1-weighted MRI in subject’s native space (**F**); Skull [11C]PBR28 TSPO PET SUV, normalized to the MNI152 space (shown as an underlay) and spatially smoothed (**G**); Reconstructed 3D rendering of the skull TSPO PET SUV in standard MNI152 space (**H**).

To standardize the PET data across all subjects, each participant’s T1-weighted MR image was registered to the MNI152 T1-weighted template using Advanced Normalization Tools (ANTs)(*32*). The resulting transformation matrices and deformation field, combined with the affine co-registration matrix mentioned above, were then applied to normalize both the skull mask and the skull PET SUV maps into the standard MNI152 space. To further minimize the potential confound of brain signals, the MNI brain template was masked out from all skull masks and skull SUV maps. To improve the signal-to-noise ratio and prepare the imaging data for voxel-wise analyses, all skull SUV PET images were smoothed using a 5-mm full-width at half maximum (FWHM) Gaussian kernel. Note that, because the brain was masked prior to spatial smoothing, this procedure did not introduce contamination of brain signal into the skull. A probability mask was subsequently generated in MNI space from the combined skull mask of all subjects and thresholded at 50%, ensuring that only voxels present within the core of the skull region were retained for final analyses. This study-specific skull template (Fig. 1D) was used as a pre-thresholding mask in all voxel-wise analyses.

### Behavioral assessment

All participants completed the Patient-Reported Outcomes Measurement Information System 29-item (PROMIS-29) questionnaire at the time of the imaging visit. This instrument was used to assess a broad range of measures, including pain-related outcomes (pain intensity and pain interference), physical function, mental health (anxiety and depression), fatigue, sleep disturbance, and social well-being. Of note, due to a slight variation in the version of the questionnaire administered, the ‘social roles’ item differed in wording across participants, i.e., “ability to participate in social roles and activities” (N=37) vs “satisfaction with social roles and activities” (N=88). Given the conceptual similarity of the two formulations, we have elected to combine the data across versions for analysis. Raw scores were used to compare groups and for regression analyses against the imaging data. For added interpretability of the PROMIS-29 scores, we also computed and reported T-scores.

### Statistical analysis

The normality of the distribution of data was assessed using the Kolmogorov-Smirnov test. Continuous variables were analyzed using the student’s t-test, and the Chi-square test of association was used to compare categorical measures between groups. As mentioned above, in our main analyses, cLBP and KOA were modeled as separate regressors, and their corresponding contrasts were averaged to yield a single ‘Pain group’ estimate representing the overall effect across both patient cohorts. For follow-up, exploratory analyses, the two pain subgroups were examined separately. For PET analyses, the following covariates were included in our models: sex and age (as informed by our preliminary regression analyses, Fig. S2) and TSPO genotype (*18, 19*) were included as covariates in the model. Statistical analyses of demographic and clinical variables were conducted using SAS version 9.4 (SAS Institute Inc, Cary, NC, USA) and R version 4.3.2. [^11^C]PBR28 PET signal was analyzed using a voxel-wise permutation-based approach(*33*) implemented in FSL’s FEAT GLM tool (www.fmrib.ox.ac.uk/fsl, version 6.0.51), using a cluster-forming threshold of z=3.1, and a cluster size significance threshold of p=0.05 to correct for multiple comparisons. In addition to the voxel-wise results (i.e., regression analyses), the following are presented for display purposes: 1) the average PET images for both patient and control groups; 2) the average PET images for patient subgroups split into tertiles in relation to PROMIS-29 pain-related scores (tertile-based cutoffs were chosen -rather than clinical thresholds or other methods- to ensure balanced signal averaging across the full range of values observed in our sample). As mentioned above, we conducted regression analyses against clinical variables only within the Pain group, and not within the healthy control group, given the limited dynamic range and overall low scores reported by the control group. These results were mapped onto a 3D rendering of the study-specific skull mask in the standard MNI152 space. Additionally, for illustrative and follow-up analyses, mean TSPO PET signal values were extracted from 5 mm-radius spheres centered on the voxel of peak significance for each cluster identified as significant in voxel-wise analyses. Each sphere was intersected with the study-specific skull mask to ensure that only local signal within the skull region was included. The extracted mean TSPO values were then visualized using box and scatter plots across the KOA, cLBP, and HC groups, where applicable. For PROMIS-29 domains, although correlation coefficients were derived from partial correlations, the plots display the actual range of scores to enhance the interpretability of the results.

## Data Availability

All data produced in the present study are available upon reasonable request to the authors.

## List of Supplementary Materials

Materials and Methods

Results

Fig. S1 to S6

## Acknowledgments

The authors would like to acknowledge Erin J. Morrissey, Angel Torrado-Carvajal, Atreyi Saha, Paulina Knight, Chelsea Pike, Ava Axelrod, Erin Hardy, Yang Lin, and Bang-Bon Koo for their help in data collection and/or interpretation of the results. Authors are also thankful to Grae Arabasz, Shirley Hsu, and the Athinoula A. Martinos Center for Biomedical Imaging Radiopharmacy team and imaging technologists for producing and administering the radioligand and assisting with the integrated PET-MR imaging.

## Author contributions

Conceptualization: MM, MLL

Methodology: MM, NE, LB, JJS, FET, MLL

Investigation: MM, LB, MPW, WCB

Visualization: MM, NE, LB, MLL

Funding acquisition: JMG, MLL

Project administration: MK, JGM, MLL

Supervision: CC, RRE, VN, JMG, MLL

Data collection and curation: MM, NE, LB, JC, MK, JPM, ZA, GG, JDH, CAC, TGC, DY

Writing – original draft: MM, NE, MK, JMG, MM

Writing – review & editing: MM, NE, LB, JC, MK, JPM, ZA, GG, JHS, CAC, TGC, DY, JJS, FET, RAC, PMG, CC, SMS, RRE, VN, KS, MN, JMG, MLL

## Data and materials availability

Deidentified data may be shared with other researchers upon reasonable request.

