## Supplementary for "Evidence for a role of skull bone marrow in human chronic pain"

### Supplementary Materials

#### Materials and Methods

##### PET-CT Acquisition

To evaluate the accuracy of pseudoCT (i.e., MRI-derived skull maps generated from T1-weighted images) for PET attenuation correction, we compared [$^{11}$C]PBR28 PET quantification using this approach against the gold-standard CT-based attenuation correction, leveraging a test dataset from a Gulf War Illness study conducted in collaboration with Boston University. In this study, adult participants (N=7, mean age: 60.22 ± 6.3 years, 100% male) underwent integrated [$^{11}$C]PBR28 PET/MRI scanning, following procedures identical to those reported in the Methods section of this manuscript (i.e., using the same scanner, radioligand, dose, and scan duration, at the same study site, etc). On a separate date, the same subjects also underwent a brain [$^{18}$F]FDG PET/CT scan at Boston Medical Center, using a GE Discovery 710 scanner. On average PET/CT and PET/MRI scans were performed 76.6 ± 86.17 days apart, with the order of acquisition varying across participants. During the PET/CT scan, participants were intravenously injected 5 mCi of [$^{18}$F]FDG and were scanned 30-minutes post-injection. While we report this information for completeness, the [$^{18}$F]FDG data are beyond the scope of the present manuscript and will not be further discussed. The CT imaging parameters included a 40 mm axial coverage and acquisition of 47 slices with a helical thickness of 3.75 mm, a display field of view (FOV) of 50 cm, tube current of 55 mAs, voltage of 120 kVp, rotation time of 0.5 seconds, pitch of 0.984:1, and table speed of 39.37 mm per rotation. The total CT acquisition time was 2.5 seconds.

In order to evaluate the accuracy of MR-based skull estimation and its impact on the reconstructed [$^{11}$C]PBR28 PET images, we reconstructed each participant's [$^{11}$C]PBR28 PET data using two different methods: 1) using the MR-based attenuation correction described in this manuscript and 2) using the CT-based attenuation correction. For the latter, we first semi-automatically removed the bed and supporting cushions from the CT images prior to use in attenuation correction. Due to the smaller axial FOV of the GE Discovery 710, the acquired CT images only partially covered the PET/MR FOV. To enable reconstruction of the full PET images with both attenuation correction methods, the missing regions in the CT images were supplemented with the corresponding regions from the pseudoCT, as illustrated in Fig. S5A. After reconstruction, the PET images obtained with both attenuation correction methods were processed following the same pipeline, masked using the bone values computed from both pseudoCT and CT ($\mu$ = 0.125 cm$^{-1}$) with an additional a 1-mm dilation applied to the head masks, to ensure complete skull coverage. The comparison between attenuation correction methods was conducted by (1) calculating the Dice similarity coefficient between the skull estimates derived from the CT and pseudoCT, and (2) performing voxel-wise correlations between the CT-based and pseudoCT-based [$^{11}$C]PBR28 PET images within the skull masks.

#### Results

##### Agreement between CT and pseudoCT

A comparison between CT and pseudoCT skull estimates, revealed that the DICE values were approximately 0.9, indicating an overall excellent agreement between predicted pseudoCT and reference CT images (Fig. S5B).

As shown in Fig. S6, voxel-wise correlations between CT-based and pseudoCT-based PET SUV within the skull masks reveal very strong linear relationships across all subjects ($r > 0.95$). Importantly, we did not see a particular pattern that would be suggestive of a systematic bias in the SUVs.

**Demographics and clinical characteristics of Pain subgroups**

Table 1 summarizes the demographic and clinical characteristics of patient subgroups.

Within the Pain group, patients with KOA (66.59 ± 8.29 years) were significantly older than those with cLBP (47.32 ± 15.45 years, $p<0.05$). KOA patients reported slightly higher pain intensity (4.32 ± 2.54) than cLBP patients (4.17 ± 1.79), though this difference was not statistically significant. In contrast, cLBP patients had worse physical function (12.64 ± 5.88) and higher depression scores (5.64 ± 2.59) compared to KOA patients (13.4 ± 3.65 and 4.97 ± 1.4, respectively; both $p<0.05$). Within the Pain Group, pain interference was rated as mild in 84 individuals (67.2%), moderate in 39 (31.2%), and severe in 2 (1.6%); anxiety symptoms were mild in 109 subjects (87.2 %), moderate in 14 (11.2%) and severe in 2 (1.6%), whereas depressive symptoms were mild in 117 participants (93.6%), and moderate in 8 (6.4%); no participants reported depressive symptoms in the severe range. Within the same group, fatigue levels were mild in 112 individuals (89.6 %) and moderate in 13 (10.4%). Regarding physical function, 27 individuals (21.6 %) reported mild, 47 (37.6%) moderate, and 20 (16%) severe difficulty. Sleep disturbance was reported as mild by 115 individuals (92 %), moderate by 9 (7.2%), and severe by 1 (0.8%). In the domain of social roles, 23 individuals (18.4 %) had mild, 31 (24.8%) had moderate, and 17 (13.6%) had severe difficulty.

Pain interference, anxiety, fatigue, sleep disturbance, and satisfaction with participation in social roles did not significantly differ between the two patient subgroups.

**TSPO PET in patient subgroups**

KOA patients demonstrated higher skull TSPO PET signal compared to cLBP patients, both within the cluster identified in the Pain>HC analyses (Fig. 2C) and in voxel-wise analyses (see Fig. S4).

#### Supplementary Figures

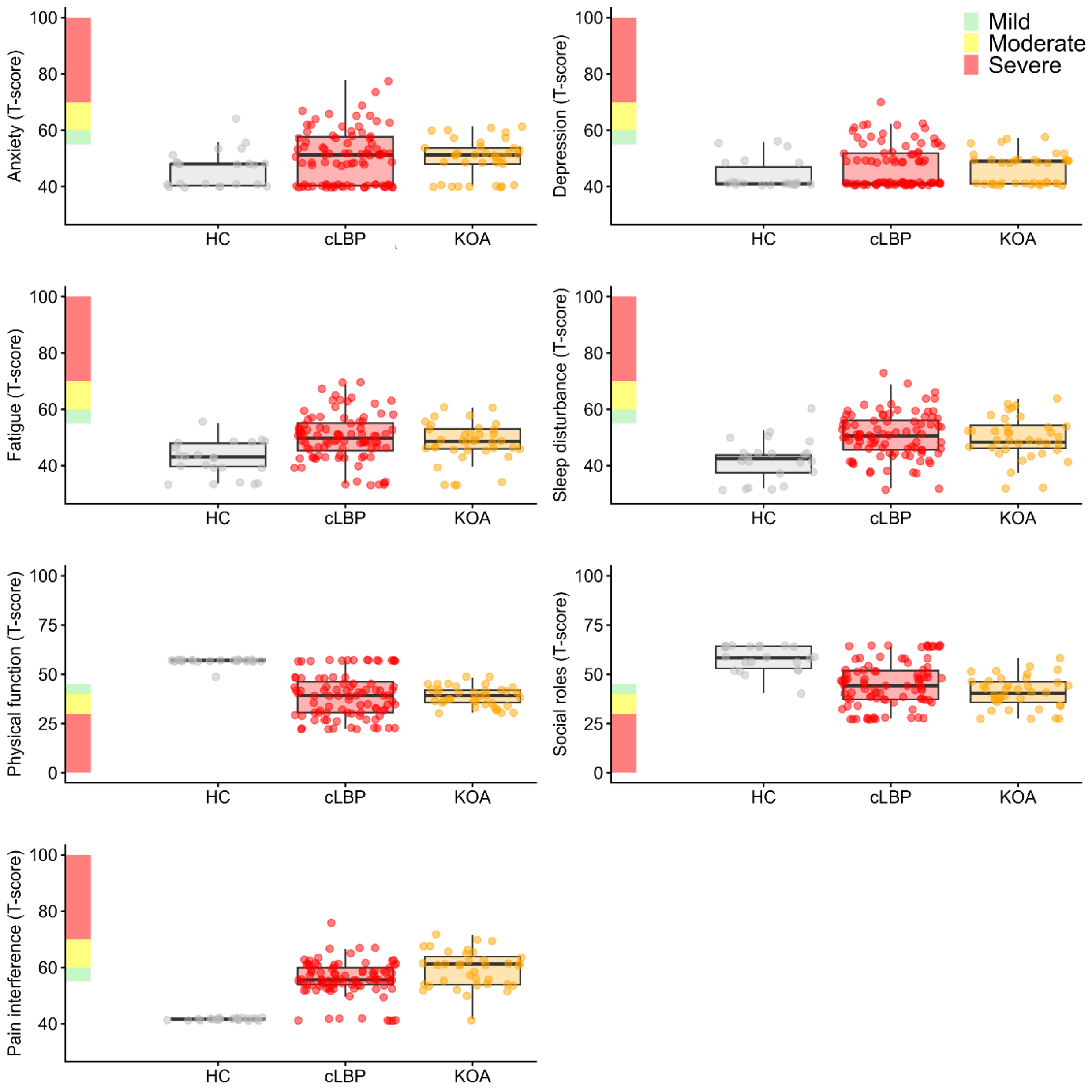


**Fig. S1. Distribution of T scores across all PROMIS 29 domains.** Distribution of the T-scores for all PROMIS-29 domains in all participants. The shaded bar on the left indicates severity thresholds: mild (green), moderate (yellow), and severe (red).

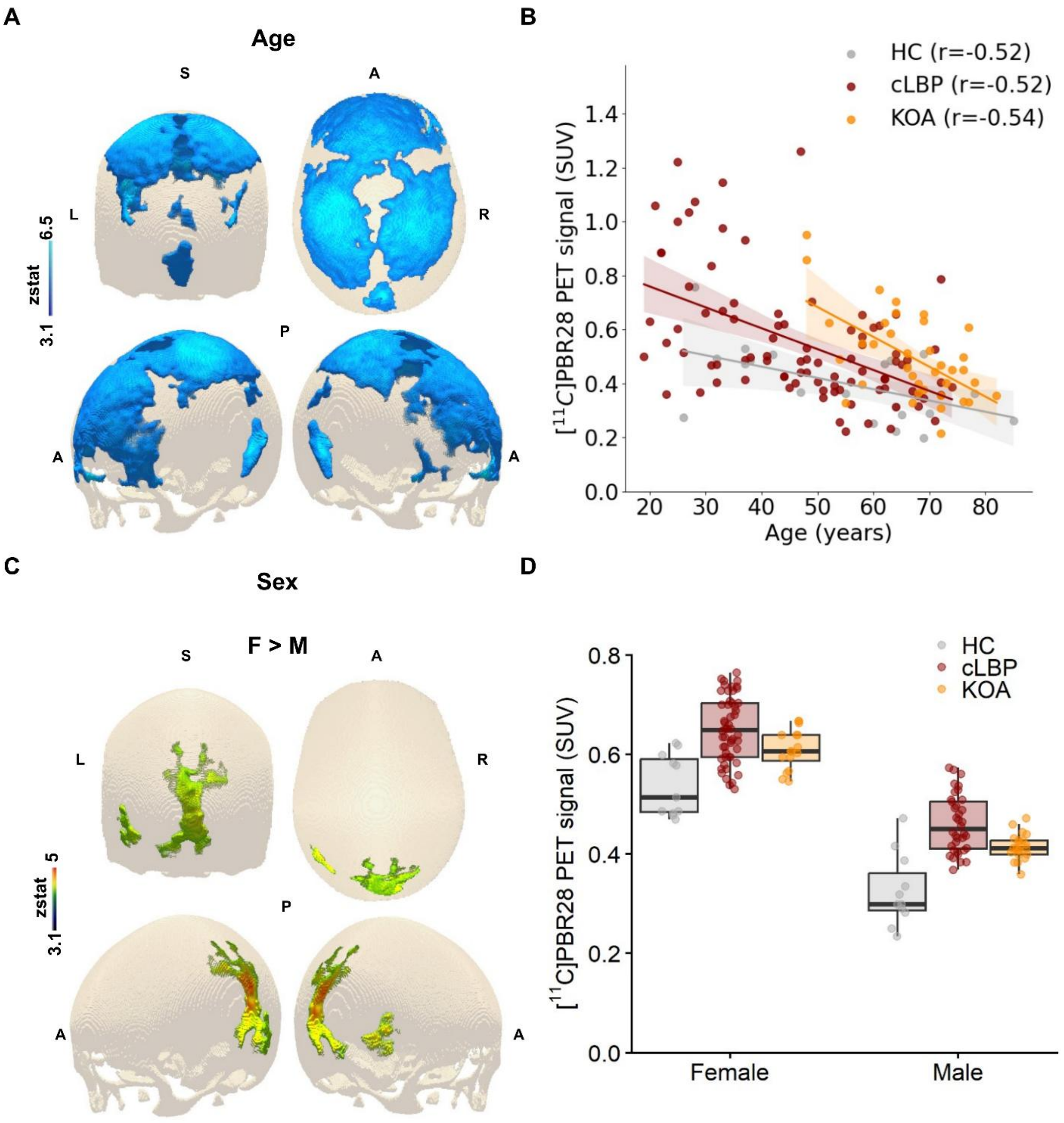


**Fig. S2. Age and sex differences in skull TSPO PET signal.** 3D rendering (**A**), and relative scatterplot (**B**), of the statistical contrast showing negative correlations between $[^{11}C]$PBR28 TSPO PET SUV and age, computed in the standard MNI152 space (**A**); 3D rendering (**C**), and relative boxplot (**D**), of the statistical contrast demonstrating higher TSPO PET signal in females compared with males. Scatter plots include the fitted regression line and 95% confidence interval of the significant voxels around the peak, averaged for each subject, by group. SUV: Standardized Uptake Value.

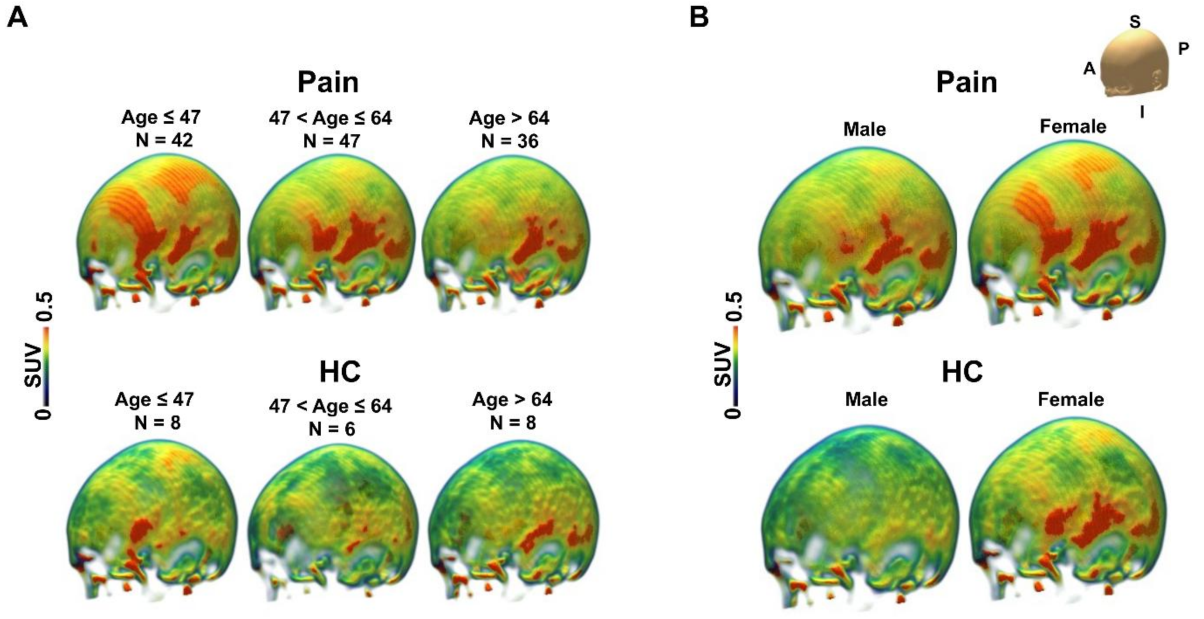


**Fig. S3. Skull TSPO PET signal by age and sex.** Average [$^{11}$C]PBR28 TSPO PET skull SUV maps by low, mid, and high tertiles of age (**A**); Average [$^{11}$C]PBR28 TSPO PET skull SUV maps by sex (**B**).

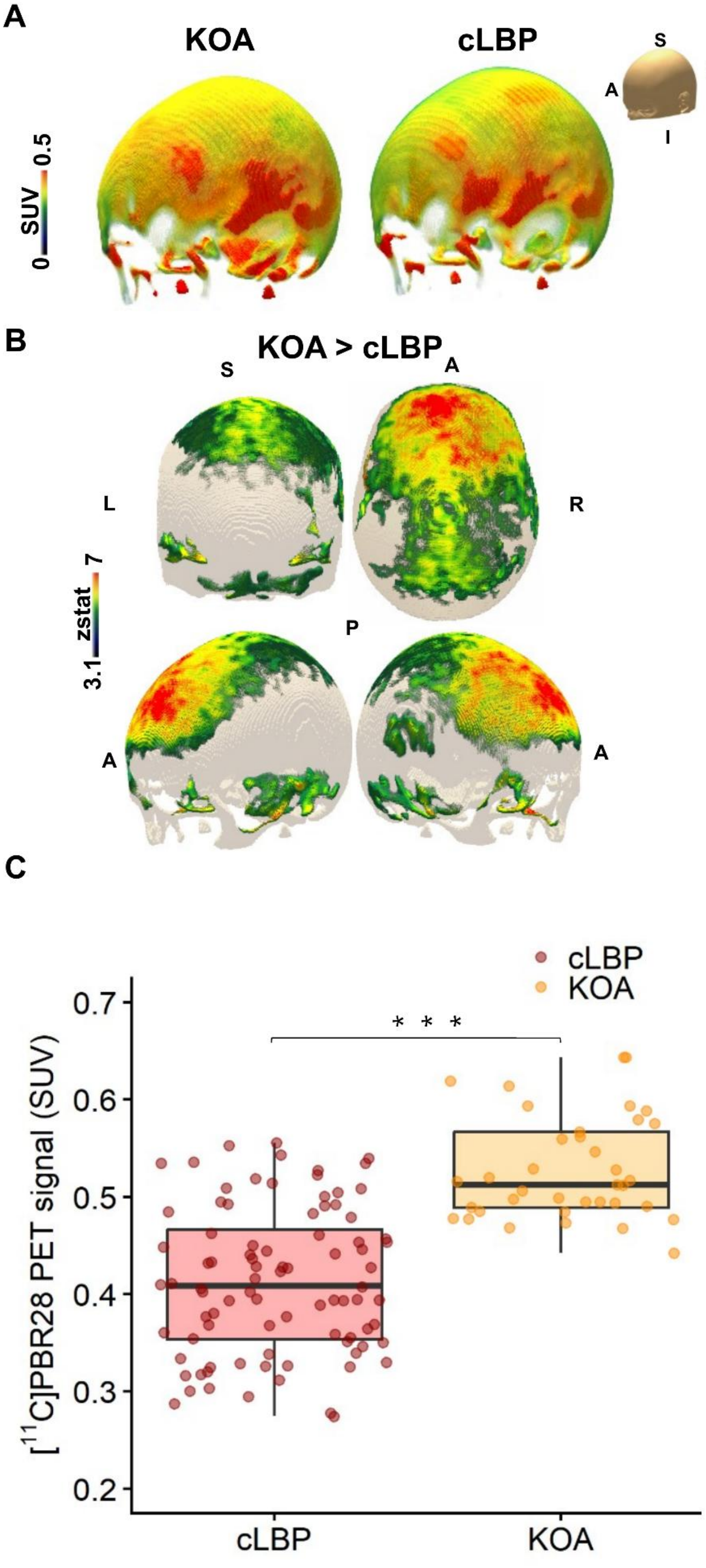
A
KOA
cLBP
S
P
A
I
SUV
0
0.5
B
KOA > cLBP
S
A
L
R
P
A
A
zstat
3.1
7
C
cLBP
KOA
* * *
[11C]PBR28 PET signal (SUV)
0.7
0.6
0.5
0.4
0.3
0.2
cLBP
KOA

**Fig. S4. Skull TSPO PET signal in chronic pain subgroups.** Average [$^{11}$C]PBR28 TSPO PET skull SUV from by Pain subgroup (KOA and cLBP) (**A**); 3D rendering (of the statistical contrast demonstrating higher TSPO PET signal in the KOA group compared with cLBP (**B**); Boxplot depicting mean TSPO PET signal from the skull region around the peak significance for back and knee pain (**C**); SUV: Standardized Uptake Value; KOA: Knee osteoarthritis; cLBP: Chronic Low Back Pain.

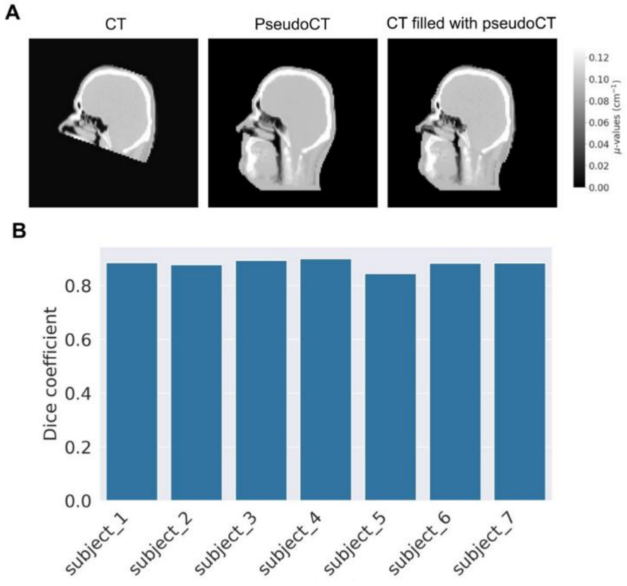


**Fig. S5. Comparison of CT and pseudoCT skull masks.** CT, pseudoCT, and filled CT with pseudoCT images of a representative subject (**A**). The corresponding DICE indices comparing the final skull masks derived from CT and pseudoCT for each subject (**B**).

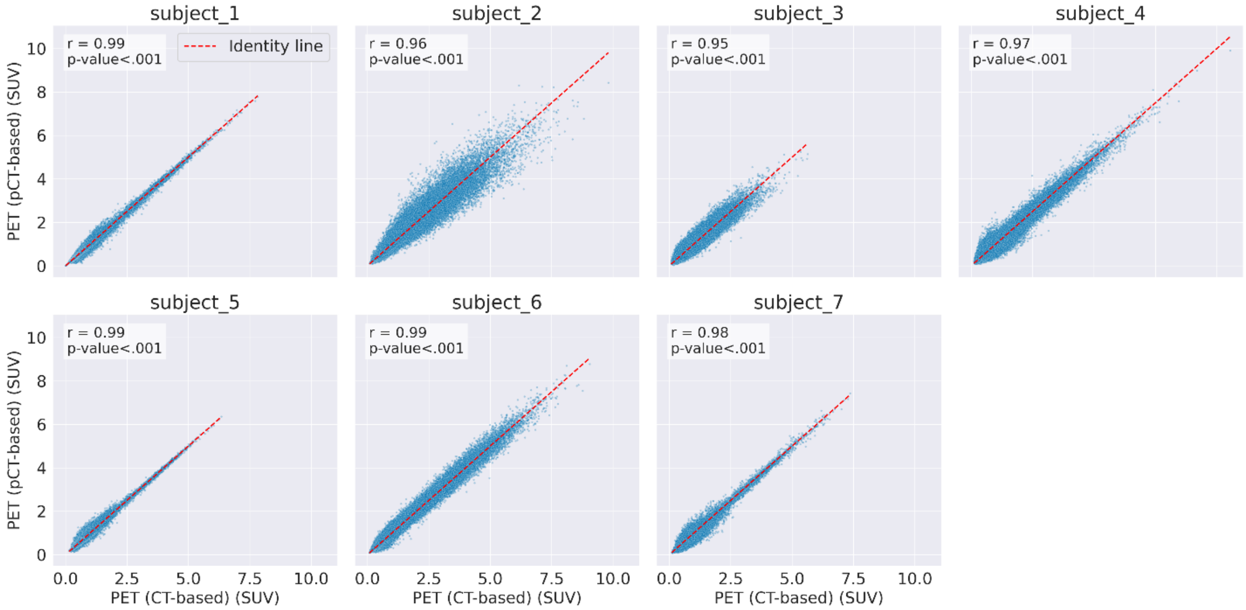


**Fig. S6. Correlation between CT and pseudoCT PET images in skull masks.** Voxel-wise correlation plots comparing CT-based and pseudoCT-based PET images within the skull masks. SUV: Standardized Uptake Value.
